# CONSORT2025: a Shiny/R application for generating CONSORT 2025-compliant flow diagram for randomized two-arm clinical trials with comprehensive data input options

**DOI:** 10.1101/2025.08.01.25332789

**Authors:** Sarowar Alom, Srimoyee Ghosh

## Abstract

Clear and accurate reporting of clinical trials is essential for transparency and reproducibility. Standardized flow diagrams are crucial for illustrating patient enrollment, allocation, follow-up, and analysis, according to the CONSORT 2025 guidelines. We created a Shiny/R web application (https://sarowar.shinyapps.io/sasg_consort/) to assist researchers by simplifying the creation of flow diagrams that comply with CONSORT and by providing a comprehensive trial data sharing option. The application offers a download option, an intuitive user interface, and automatic diagram generation for structured data input. Both clinical researchers and trial coordinators, particularly those without programming experience, can use it. Our tool supports the broader objectives of enhancing uniformity and transparency in clinical research reporting.

## 1. Background

The integrity, reproducibility, and application of biomedical research findings depend on the accurate and transparent reporting of clinical trials. The Consolidated Standards of Reporting Trials (CONSORT) Statement, first published in 1996 and revised subsequently, provides structured guidelines for reporting randomized controlled trials (RCTs) [1–3]. In 2025, the CONSORT guidelines were revised to incorporate the recent developments in trial methodology and reporting practices, with a thorough focus on participant flow reporting, documentation of protocol deviations, and detailed analytical transparency [1].

Even though journals and regulatory agencies have widely endorsed CONSORT recommendations, many investigators still struggle to accurately and consistently detail the participant flow diagram, which is a crucial part of the CONSORT checklist [4]. On many occasions, the CONSORT diagram lacks key information such as reasons for lost to follow-up, reasons for not receiving allocated intervention and reasons for exclusion of some participants [5]. This information is necessary for the readers to understand the trial. Although R packages have been developed to assist in constructing CONSORT flow diagrams, their use generally requires prior knowledge of the R programming language, which limits accessibility for researchers.

To address these challenges, we developed an interactive web-based application utilizing the R Shiny framework, designed to facilitate the generation of a CONSORT 2025-compliant participant flow diagram for randomized two-arm clinical trials. The application enables users to systematically input trial data through a guided interface, automatically updating the standardized flow diagram in real-time. Furthermore, users can enter a detailed record of the data concisely represented in the CONSORT flow diagram as the trial transparency information. Both the flow diagram and the trial transparency information can be downloaded in the form of HTML files and can be converted to any file type according to the user’s need. The trial transparency information can be submitted to external repositories, such as Zenodo or Figshare, and links can be shared. Therefore, we aim to enhance adherence to reporting standards, improve the quality of trial reporting, and ultimately contribute to greater transparency and reproducibility in clinical research.

## 2. Methods

The project had the following objectives:

1. To develop the Shiny R application for the generation of a CONSORT flow diagram in compliance with the latest CONSORT guideline [1].
2. To provide users with a space to systematically incorporate trial data as trial transparency information for a better understanding of the participant flow.

The project was written and established in R (version 4.3.3) using multiple R packages, and the shiny application was submitted to Shiny server (https://sarowar.shinyapps.io/sasg_consort/). Extensive testing and validation were done using mock datasets representing randomized two-arm clinical trials.

## 3. Results

The CONSORT Shiny/R application link takes the user to the landing page as shown in Fig. 1. In the data entry panel, users can enter the data manually and the flow diagram updates in real time. At the bottom of the data entry panel, the HTML download option is available. This download option exports the flow diagram in HTML format (Fig. 2). The right side panel is the trial transparency information data entry panel. In this panel, users can enter detailed information about the participant flow, concisely described in the flow diagram. The information provided in the trial transparency panel can also be downloaded in HTML format (Fig. 3). We encourage users to extensively use the trial transparency panel and share the data along with the CONSORT flow diagram.

**Fig. 1.**
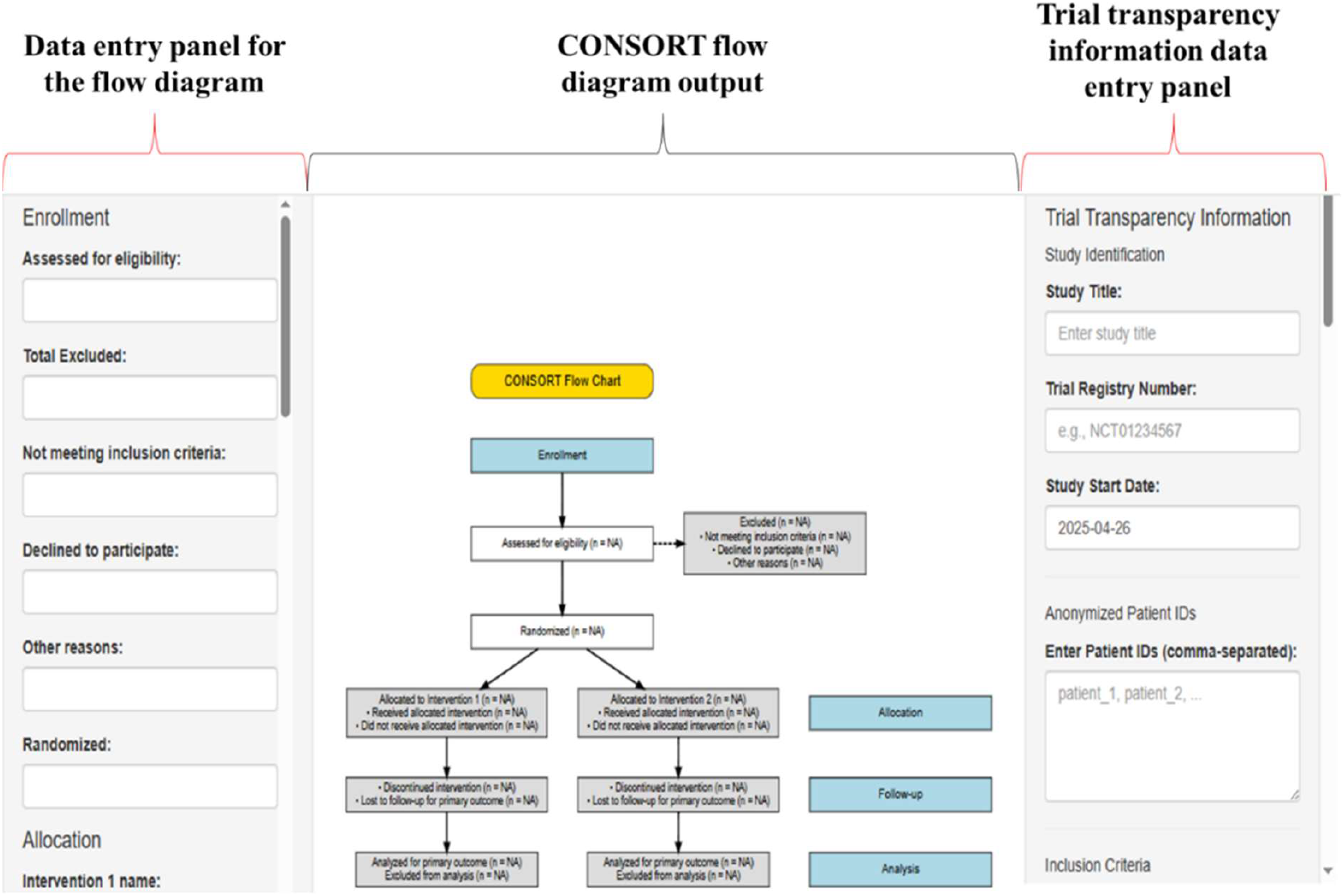
CONSORT2025 app landing page.

**Fig. 2.**
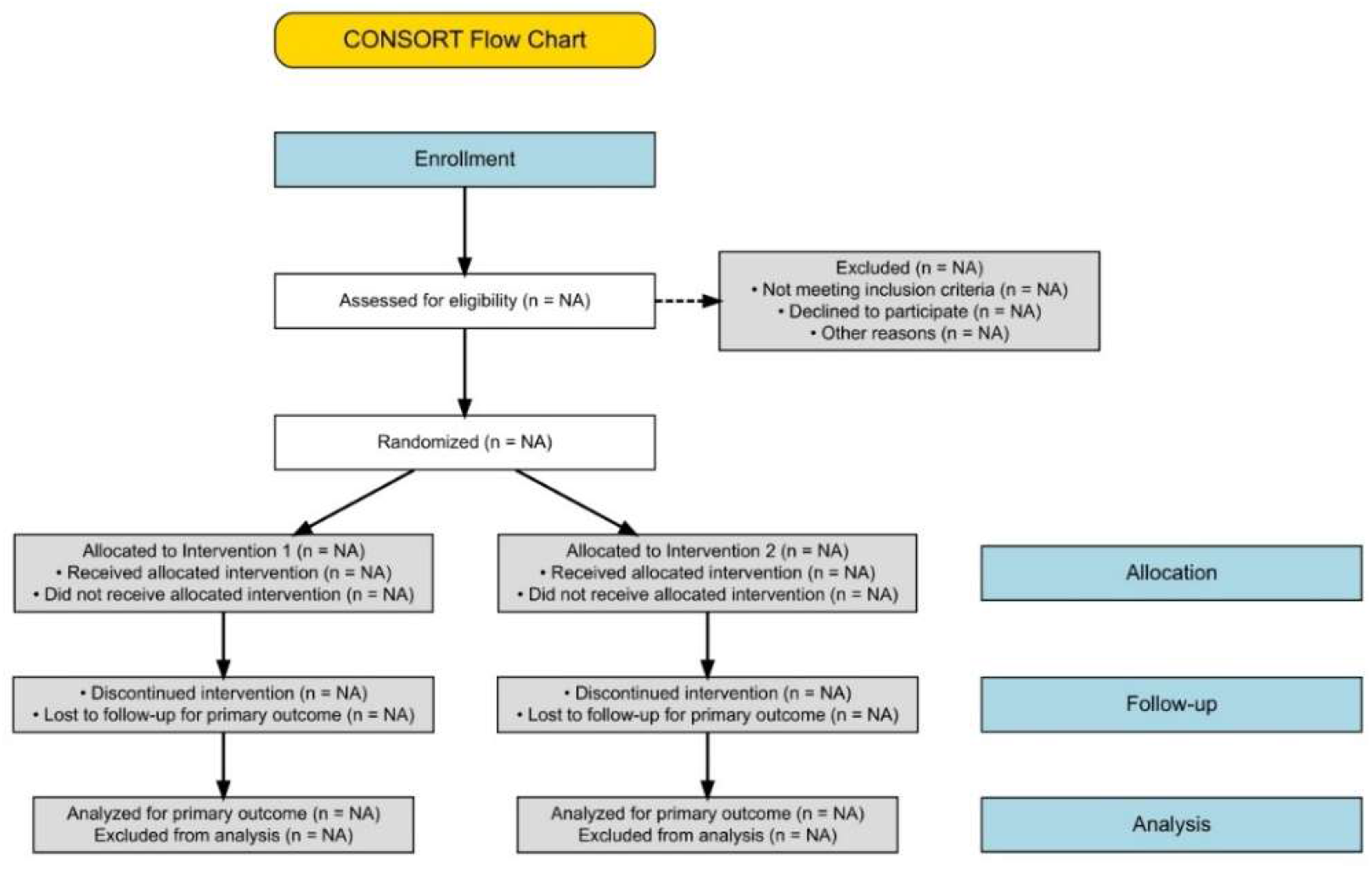
Representation of the CONSORT flow diagram generated using the Shiny/R application.

**Fig. 3.**
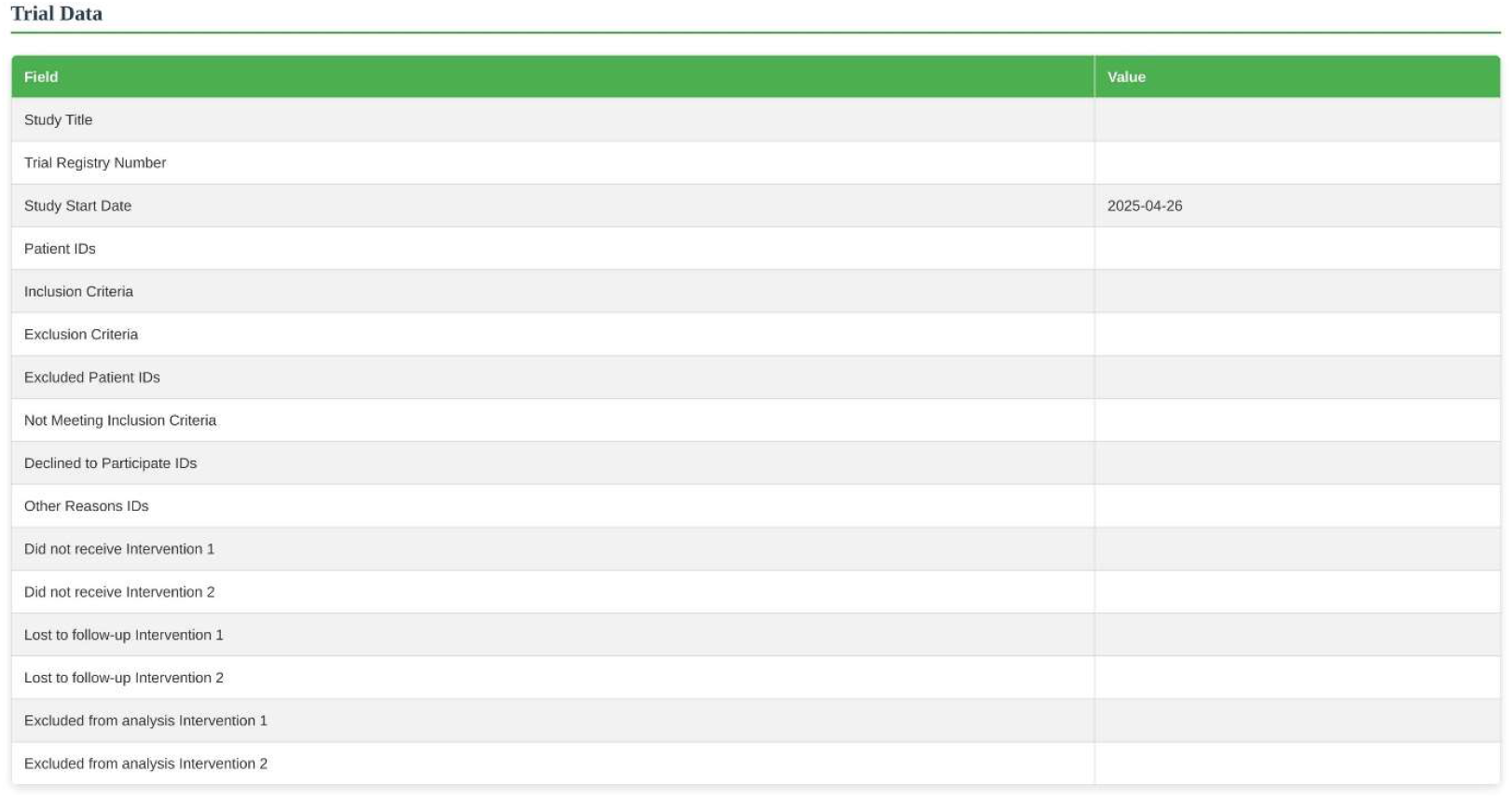
Representation of the trial transparency information generated using the Shiny/R application.

## 4. Discussion and conclusion

Our Shiny R web application was developed to make it easier for researchers to create CONSORT flow diagrams for clinical trials and also provide space to share the patient flow data in detail. Following the updated CONSORT 2025 guidelines [1], clear reporting of trial progress is very important to improve transparency and trust in research. However, CONSORT diagrams lacking key information make it difficult for readers to understand the trial.

Our application provides a simple and user-friendly interface where users can input trial data and instantly see a preview of the CONSORT diagram. It also offers downloads in HTML, making it easy to use the diagrams in journal submissions or presentations.

## Data Availability

All data produced in the present work are contained in the manuscript

